# A single dose of lamotrigine induces a positive memory bias in healthy volunteers

**DOI:** 10.1101/2023.12.06.23299597

**Authors:** Tarek Zghoul, Pilar Artiach Hortelano, Alexander Kaltenboeck, Lucy Wright, Guy Goodwin, Liliana Capitão, Catherine Harmer

## Abstract

**Background:** Lamotrigine has been shown to be effective in the long-term treatment and relapse prevention of depression in bipolar disorder. However, the neuropsychological mechanisms underlying these effects are unclear. We investigated the effects of lamotrigine on a battery of emotional processing tasks, previously shown to be sensitive to antidepressant drug action in healthy volunteers.

**Methods:** Healthy volunteers (n=36) were randomised in a double blind design to receive a single dose of placebo or 300mg lamotrigine. Mood and subjective effects were monitored throughout the study period and emotional processing was assessed using the Oxford Emotional Test Battery (ETB) 3 hours post administration.

**Results:** Participants receiving lamotrigine showed increased accuracy for the recall of positive vs. negative self-descriptors, compared to those in the placebo group. There were no other signficant effects on emotional processing in the ETB, and lamotrigine did not affect ratings of mood or subjective experience.

**Conclusions:** Lamotrigine induced a positive bias in emotional memory, similar to the effects of other antidepressants as reported in previous studies. Further work is needed to assess whether similar effects are seen in the clinical treatment of patients with bipolar disorder and the extent to which this is associated with its clinical action in relapse prevention.

## Introduction

Bipolar disorder (BD) is characterised by recurrent presentations of elevated mood episodes, in the form of either mania or hypomania, and depressive episodes (APA, 2022). Affecting over 1% of the global population, this severe chronic affective illness is among the leading causes of worldwide disability (Global Burden of Disease Study, 2015; Judd et al., 2002). BD’s predominant mood state is bipolar depression (Malhi et al., 2003). However, most available treatments for BD such as mood stabilisers (i.e., lithium), antiepileptic drugs (i.e., carbamazepine and valproate sodium) or atypical antiepileptic drugs (i.e., olanzapine) exert their effects primarily on mania/hypomania and, to a lesser extent, on depression (Goldsmith et al., 2003).

Lamotrigine is a relatively new drug that is associated with prevention of depressive episodes within the clinical presentation of BD. First licensed as an add-on treatment for epilepsy, lamotrigine was quickly seen to have beneficial effects in BD, gaining approval in 2003 as a maintenance treatment for BD by the US FDA (Goldsmith et al., 2003; Vajda et al., 2013). Lamotrigine inhibits voltage-gated sodium channels, and thereby reduces presynaptic glutamate release (Ketter et al., 2003; Lee et al., 2008; Corina Prica et al., 2008; Sills & Rogawski, 2020). Interestingly, the glutamatergic system has received increased attention in the pathophysiology and treatment of mood disorders in recent years. In patients with BD, altered levels of glutamate have been found in the frontal cortex, among other regions, together with alterations in the expression or function of NMDA receptor subunits (Murrough et al., 2017; Sanacora et al., 2008). These glutamatergic changes are suggested to contribute to cognitive changes seen in BD in line with glutamate’s role in synaptic plasticity, learning and memory (Sanacora et al., 2008).

BD is associated with impairments in cognition, and more specifically emotional cognition (Bogie et al., 2019; Fijtman et al., 2020). Patients with BD depression and unipolar depression show a more general impairment in recall and recognition processes, which seem to be related to poor encoding rather than to rapid forgetting (Bearden et al., 2006; Deckersbach et al., 2004). Regarding emotional cognition, the typology of affective bias in BD is complex (Bilderbeck et al., 2016), but there is evidence suggesting that these biases are mood-dependent. In patients with BD with manic symptomatology, more positive information (i.e., positive bias) is preferred, whereas those with depressive symptomatology show a negative bias in emotional processing (Paykel, 1987). Consequently, patients with BD depression showed a similar negative bias to those with unipolar depression in the categorisation of ambiguous facial expressions and self-referential emotional words (Bilderbeck et al., 2016; Godlewska & Harmer, 2021). Despite this evidence, little research has investigated lamotrigine’s effect on emotional processing and memory biases in BD. A study involving a paedriatic BD sample found that lamotrigine improved memory performance, which could be related to its glutamatergic properties (Pavuluri et al., 2010). Interestingly, in a naturalistic study looking at the association between mood state and emotional bias in BD, current medication use, including anticonvulsants, was associated with a greater positive bias compared to medication-free patients (Bilderbeck et al., 2016). Although lamotrigine was not investigated specifically, it suggests that mood-stabilising treatment in BD may be associated with positive emotional bias. Evidence of the effect of lamotrigine on cognitive processes, including emotional proceses is still incomplete. Furthermore, whether the effects of lamotrigine are a consequence of symptom improvement over time or whether they represent a key mechanism underlying the action of lamotrigine is still unclear. In addition, the lack of a placebo control in previous studies, together with the use of highly heterogeneous clinical populations, limits the interpretation of such results.

Experimental medicine may represent a useful approach in the investigation of lamotrigine’s mechanism of action early within treatment, before changes in symptomatology are typically seen. The use of this approach in healthy volunteers has proven useful in characterising early effects of conventional antidepressants. Accordingly, an increase in the relative processing of positive rather than negative emotional information has been observed in healthy volunteers following single or repeated antidepressant administration from a variety of pharmacological classes (e.g., selective serotonin reuptake inhibitors, atypical antidepressants) (Harmer et al., 2017). For example, SSRIs increase the recognition of positive facial expressions in a recognition task and increase recall for positive over negative information in a free recall test. Interestingly, similar effects have also been found in depressed individuals after a single dose of reboxetine compared to placebo (Harmer et al., 2009). Ultimately, these studies have highlighted that conventional antidepressants may act by reducing negative biases characteristic of depression (Godlewska & Harmer, 2021; Harmer, 2010; Harmer et al., 2009), with these effects, which are seen rapidly and in the absence of subjective mood changes, contributing to subsequent clinical changes in mood (Harmer et al., 2017; Harmer et al., 2009).

The current study therefore aimed to use an experimental medicine approach to investigate the effects of a single dose of the novel drug lamotrigine in a healthy volunteer sample with the use of a well validated task battery measuring emotional cognition. The use of a healthy volunteer sample can be advantageous as it allows for the direct assessment of lamotrigine on neuropsychological mechansims unconfounded by disorder-related factors. Additionally, the Emotional Test Battery (ETB) has been extensively used in experimental medicine approaches to characterise different antidepressant drug classes both in healthy volunteers and clinical populations. Given that lamotrigine’s predominant effects are on BD depression and that emotional processing biases have been reported in these depressive states, this research can aid the understanding and identification of surrogate markers for improved targets of depression in BD. It was hypothesised that lamotrigine would induce a positive bias in emotional processing and cognition, with these effects being more pronounced on those tasks including a memory component as explored above.

## Material and methods

### Study sample

Thirty-six healthy volunteers (12 women and 24 men) aged 18-40 years were recruited via print and online announcements and enrolled in this study. All participants gave written informed consent prior to participation in the study and were reimbursed for their time and any other expenses. Participants completed a screening visit to ascertain eligibility prior to proceeding to the experimental session. This included a psychiatric interview (Structured Clinical Interview for DSM-5; SCID-5) and a general medical screening, with females testing negative on a pregnancy test. Participants were then assessed for estimated intelligence using the National Adult Reading Test (Revised Version, NART-R) (Nelson & Willison, 1991), trait anxiety using the Spielberger’s State-Trait Anxiety Inventory (STAI) (Spielberger, 1983), depressive symptoms using the Beck Depression Inventory (BDI) (Beck et al., 1961), and personality traits using the Eysenck Personality Questionnaire (EPQ) (Eysenck, 1975). Participants were excluded if they had (a) current or past history of psychiatric disorders (as assessed using the SCID-5), (b) insufficient English language skills, (c) current or past history of drug or alcohol dependency, (d) significant medical conditions, (e) previous participation in studies using the same emotional processing tasks, (f) contraindication to MRI, (g) history of recurrent allergies and rashes, (h) consumed any psychoactive medication or folic acid supplements, (I) smoked in excess of five cigarettes a day, (j) had a high intake of caffeinated drinks (>5 cups a day), (k) were pregnant or lactating women or (l) were taking the contraceptive pill. Ethical approval was obtained from the Central University Research Ethics Committee (CUREC). The study was registered on ClinicalTrials.gov (Identifier: NCT04396938).

### Procedure

In this double-blind, placebo-controlled, randomised, parallel group study participants were assigned to receive either a single oral dose of lamotrigine (300 mg) or placebo. All tablets were encapsulated to provide uniform appearance and ensure a double-blinded study. The randomisation code was created by another member of the lab group not involved in the experiment. Participants were asked to avoid having any alcohol for at least 12 hours and food and caffeine for at least three hours prior to the experimental visit. As lamotrigine peak plasma concentrations (T_max_) occur between one to three hours following oral intake (Goldsmith et al., 2003), all volunteers were tested three hours after drug administration. During the first two hours post drug administration, participants stayed in a quiet clinic room, where they were free to relax or engage in any activity such as reading or using their personal electronic devices. During the next hour, participants underwent a functional magnetic resonance imaging scan (results reported elsewhere), and were tested with the Oxford Emotional Test Battery (ETB) (Harmer et al., 2009) (Schmidt et al., 2015).

### Measures

#### Demographic and questionnaire measures

Self-reported questionnaires were administered to assess subjective state and side-effects at three time points: prior to active drug or placebo intake (Baseline or T1), after two hours (T2), and after three hours (T3). The following measures were used: the STAI-State (Spielberger, 1983), Positive and Negative

Affect Schedule (PANAS) (Watson et al., 1988), and visual analogue scales (VAS) of subjectively experienced emotions. Side effects/somatic symptoms were measured on 11 symptoms (agitation, dry mouth, nausea, aggression, tremor, headache, drowsiness, dizziness, back or joint pain, vision impairments and rashes) with the following scale: 0=absent, 1=mild, 2=moderate, and 3=severe.

#### Emotional test battery

The ETB is a validated tool of five broadly computerised behavioural tasks that allows for the assessment of emotional information processing in different cognitive domains (Harmer et al., 2009), and proved to be sensitive to the early effects of antidepressant medication (Harmer et al., 2009; Warren et al., 2019). With a duration of 45 minutes, it was completed in the following order: facial expression recognition task (FERT), emotional categorization task (ECAT), attentional dot-probe task, emotional recall task (EREC), and emotional recognition memory task (EMEM).

In the FERT, participants were presented with pictures of facial expressions displaying one of the six basic emotions (i.e., anger, disgust, fear, happiness, sadness and surprise) or neutral. Each emotion was displayed at 10 morphed intensity levels from “neutral” (i.e. 0%) to “full intensity” (i.e. 100%) resulting in a total of 250 pictures (Young et al., 1997). Facial expressions were presented randomly for 500 ms each, followed by a blank black screen. Participants were instructed to identify each emotion as quickly and accurately as possible by pressing one of the seven correspondingly labelled keys on the keyboard. Main outcomes were number of misclassifications, accuracy and reaction time (RT).

In the ECAT, participants were presented with 60 words representing personality descriptors that were either agreeable (e.g. honest, thoughtful, etc; *N*=30) or disagreeable (e.g. untidy, bossy, etc; *N*=30). Each word was randomly presented in the centre of a computer screen for 500ms. Participants were asked to imagine overhearing someone describing them with each word displayed and to indicate as quickly and accurately as possible whether they would like or dislike to be described as such by pressing the correspondingly labelled key on the keyboard. The main outcome measures were RT and classifications.

In the attentional dot-probe task (Schmidt et al., 2015), participants were presented with 60 positive and 60 negative words paired with neutral words. In each trial, a fixation cross was presented in the centre of the screen for 500 ms, followed by two words displayed at the bottom and top of the screen. These word pairs were shown for either 500 ms in the unmasked condition or 17 ms in the masked condition, and were followed by a mask for 483 ms. Masks were created from letters, non-letter symbols and digits and were matched in word length and position. A dot-probe (one or two stars) then replaced the masks or words in their location. Accordingly, the dot-probe could appear behind the emotional word (congruent trials) or the neutral word (incongruent trial). Participants were asked to identify the probe as accurately and quickly as possible by pressing the corresponding key on the keyboard. There was a total of 180 trials (30 negative-neutral, 30 positive-neutral and 30 neutral-neutral pairs each for the unmasked and masked conditions) with emotional words displayed at equal frequency at the bottom and top of the screen. Accuracy and RT were noted. Attentional vigilance scores were then calculated by subtracting RT in congruent trials from RT in incongruent trials (Attentional vigilance = RT incongruent trials – RT congruent trials). This task was completed in between the ECAT and the EREC.

The EREC took place approximately 15 minutes after the ECAT, from which participants were asked to write down as many words as they could recall in four minutes. The outcome measure was numbers of correctly and incorrectly recalled words.

In the EMEM, positive and negative personality descriptors (in equal frequency) were presented in the center of the screen for 500ms. In total, 120 words were randomly presented with 60 being familiar and having already been displayed during ECAT and 60 being novel or unfamiliar. Participants were asked to indicate as accurately and quickly as possible, whether the word displayed was familiar or novel by pressing the corresponding key on the keyboard. Main outcomes included RT, correct responses and false positives.

### Statistics

Analyses were performed using SPSS (version 25.0) with significant levels set at p<0.05. One participant was excluded from the attentional dot-probe task analysis as there was a computer failure during this task. Changes in subjective ratings (STAI-State, PANAS, VAS) and side effect symptoms were analysed using mixed analysis of variance (ANOVA), with timepoint (baseline or T1, T2 and T3) as within-subjects factor, and treatment group (lamotrigine or placebo) as between-subjects factor. ETB data was analysed using mixed ANOVA with emotion/valence as within-subject factors and treatment group (lamotrigine or placebo) as between-subject factor. The attentional dot-probe task was analysed using mixed ANOVA with valence and masking as within subject factors and treatment group as between-subject factor. Significant interactions were further explored using independent sample t-tests and/or paired-samples t-tests. Whenever relevant, analyses were repeated with side-effects as a covariate (averaged values for timepoint 3).

## Results

### Baseline measures

Sample’s baseline characteristics can be found in Table 1. Participants were well matched on age, gender distribution, proportion of university educated participants, ethnicity, verbal IQ, the BDI and the EPQ scales. However, the lamotrigine group displayed higher scores on the STAI-Trait.

**Table 1.** Baseline characteristics of the sample.

|  | Placebo (n=18) | Lamotrigine (n=18) |
| --- | --- | --- |
| *Demographics* |  |  |
| Gender, n |  |  |
| Male | 12 | 12 |
| Female | 6 | 6 |
| Age, years | 24.39 (4.29) | 23.83 (4.67) |
| Education |  |  |
| University | 16 | 17 |
| Non-University | 2 | 1 |
| Ethnic background |  |  |
| White | 12 | 12 |
| Asian/Asian British | 5 | 4 |
| Black/African/Caribbean/Black British | 1 | 0 |
| Mixed/Multiple ethnic groups | 0 | 2 |
| *Baseline clinical measures* |  |  |
| NART | 118.72 (5.29) | 118.67 (4.37) |
| STAI-trait | 29.61 (5.12) | 33.44 (4.95) |
| BDI | 1.89 (1.88) | 2.72 (3.10) |
| EPQ-neuroticism | 4.82 (2.92) | 7.33 (4.96) |
| EPQ-psychoticism | 2.53 (2.4) | 3.22 (1.63) |
| EPQ-Lie/Social Desirability | 8.65 (3.35) | 9.72 (4.54) |
| EPQ-Extraversion | 16.12 (3.84) | 13.61 (5.67) |

Data given as mean (standard deviation) unless otherwise indicated. N, sample; NART, National Adult Reading Test; BDI, Beck Depression Inventory; STAI, State-Trait Anxiety Inventory; EPQ, Eysenck Personality Questionnaire.

### Changes in subjective ratings and side effects

Measures of subjective mood were administered to assess subjective state and somatic side effects following treatment administration. There were no significant effects of treatment on the PANAS, VAS nor the STAI-State (all *p*>0.05). There was a significant main effect of group on side effects with the lamotrigine group reporting more side effects across all time points, including more physical symptoms at baseline (F(1.34)=6.967, *p*=0.012). There was no significant interaction between group and time points (p=1.19).

### FERT

For the accuracy at recognizing facial expressions’ emotions and number of misclassifications there were no significant main effects of treatment nor significant interactions between treatment and emotions (all F_s_<6, all *p*_s_>0.12).

### ECAT

For accuracy of the emotional recall and RTs, there were no significant main effects of treatment nor significant interactions between treatment and emotion (all F_s_<1, all *p*_s_>0.31).

### Attentional Dot-Probe task

For vigilance scores during the attentional dot-probe task, there was no significant main effect or interaction with treatment (p>0.10).

### EREC

For accuracy rates, there was a significant interaction between group and emotion (F(1,34)=4.953, p=0.03). This was driven by the lamotrigine group accurately recalling a significantly higher proportion of positive (8.78) vs negative words (6.44) compared to placebo (F(1,17)=14.875, *p*=0.001, Figure 1). Repeating this analysis with side-effects as a covariate yielded a similar pattern of results (F(1,33)=5.188, p=0.03).

**Fig. 1.**
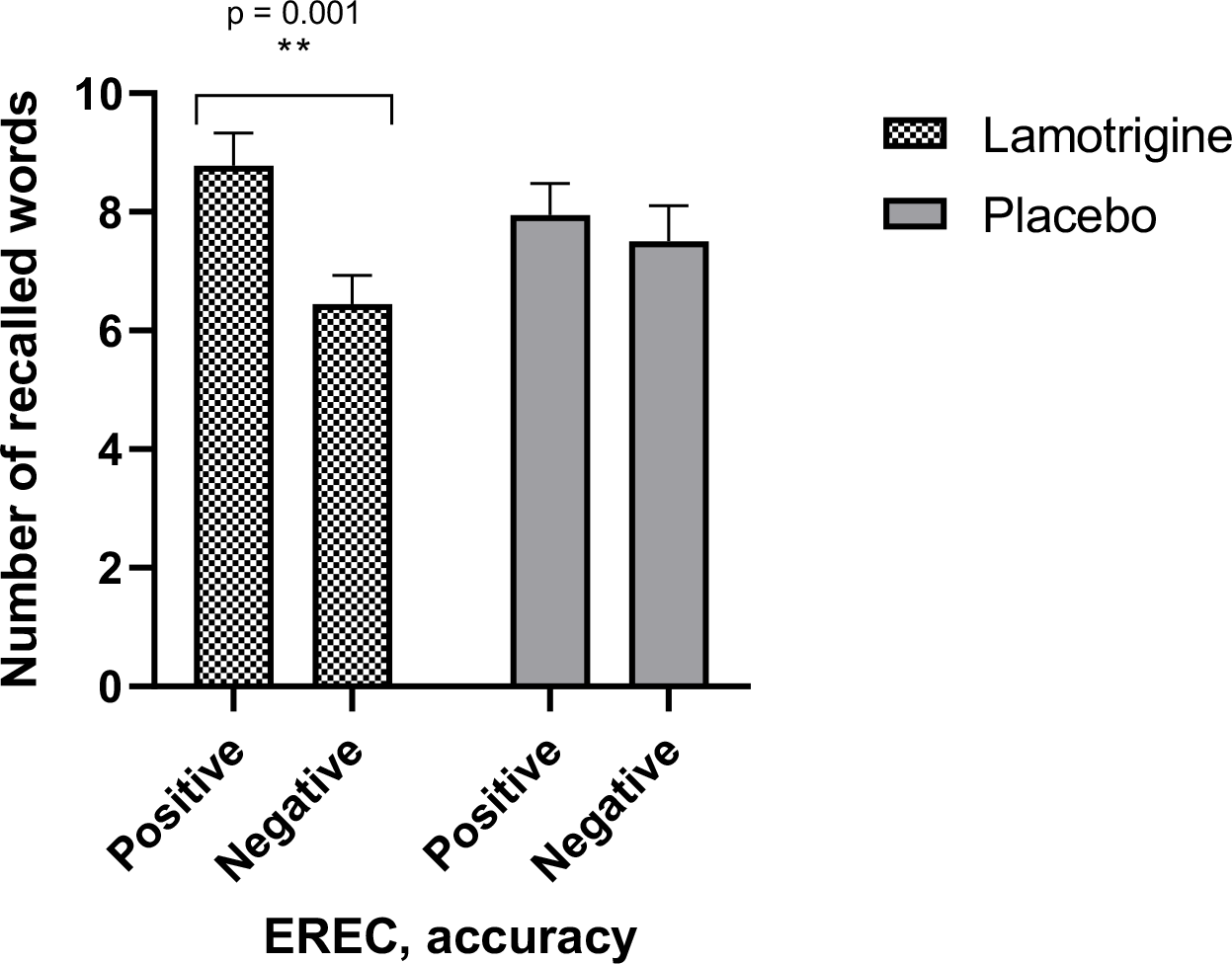
Accuracy of positive vs negative recalled words between treatment groups. Comparison between the number of recalled words (mean) for positively vs negatively valenced words in participants given lamotrigine vs placebo. Bars represent the standard error of the mean. Asterisks represent a statistically significant difference between positive and negative words.

### EMEM

For accuracy rates and RTs of correctly recalled words there were no significant main effects of treatment and no significant interaction between group and emotion (all F_s_<1.34, all *p*_s_>0.40).

## Discussion

The present study investigated the effects of a single oral dose of lamotrigine (300mg) on emotional cognition in a healthy volunteer sample following a randomised double-blind, parallel placebo-controlled design. As such, this study aimed to characterise the neuropsychological mechanisms underlying lamotrigine, to ultimately help enlighten how lamotrigine is beneficial in the prevention of depressive episodes in BD. Three hours after the treatment administration, the lamotrigine group had a significant higher accuracy for the recall of positive vs. negative words compared to placebo. There were no other statistically significant effects in the analysis of the remaining ETB measures. These results are in line with a positive bias induction in emotional memory following acute lamotrigine treatment. There were differences between groups in subjective ratings of physical symptoms but these were also seen before administration of lamotrigine and most likely reflect a naturally occurring difficulty of group matching via randomisation. Controlling for these differences statisticially did not affect the positive bias in emotional memory seen following lamotrigine.

### Lamotrigine induced a positive bias in memory recall

Participants in the lamotrigine group significantly recalled more positive than negative self-referential words in the emotional recall task, compared to those in the placebo group. This positive shift in memory has been extensively reported in experimental medicine studies of conventional antidepressant drugs using healthy volunteers. For instance, acute administration of mirtazapine (Arnone et al., 2009) or reboxetine, and 7-day treatment with citalopram or reboxetine increased positive affective memory in healthy participants (Harmer et al., 2017). Preclinical models of affective bias have also revealed early changes in negative memory biases with a range of treatments used in depression (Stuart et al., 2017). These positive shifts on emotional cognition have been proposed as a working mechanism behind antidepressant drug action (Harmer et al., 2017), and may be key in lamotrigine’s effectiveness in preventing BD depression.

Biases affecting emotion and cognition in BD have been reported, with these being mood-dependent (Bilderbeck et al., 2016). Accordingly, a mood-congruent schema affecting attention, interpretation and memories about the world, oneself and the future has been proposed. In patients with BD with depressive symptomatology, negative information is preferred (i.e., negative bias); whereas in those with manic symptomatology, more positive information is favoured (i.e., positive bias) (Paykel, 1987). Furthermore, a negative cognitive style in people with BD has been reported to be a vulnerability factor that, when activated, triggers a cycle of rumination and negative self-related thoughts that feed into depression (Hitchcock et al., 2020; Van der Gucht et al., 2009). In line with this negative bias, greater instability of depressive symptomatology in patients with BD correlated with negative biases in the categorisation of emotional of self-referential emotional words (Bilderbeck et al., 2016; Godlewska & Harmer, 2021). Further emotional memory biases in BD have been observed, with patients showing enhanced memory for content that is associated with adverse emotions and for neutral content that preceded the emotional stimulus (Bogie et al., 2019; Fijtman et al., 2020). Additionally, both in BD depression and unipolar depression, patients show a worse recall for positive autobiographical memories, where these are less specific and more categorical, compared to healthy controls (Young et al., 2016). Understanding how these affective and cognitive biases in BD interact with both disorder stages and treatment can be helpful to determine treatment efficacy, and explore new potential treatments for BD. Consistent with previous research on conventional antidepressants (Harmer et al., 2017), lamotrigine may be shifting depressive-related negative biases into more positive ones. These effects may also help stabilise mood and prevent the development of depressive symptomatology in BD.

Lamotrigine’s effect on emotional memory might be related to its effects on glutamate neurotransmission (Lee et al., 2008; C. Prica et al., 2008; Sills & Rogawski, 2020). With a key role in the regulation of synaptic plasticity, the glutamatergic system is involved in processes such as learning and memory, among others (Murrough et al., 2017). Both animal and human studies have reported a clear effect of lamotrigine on memory. In mice, lamotrigine administration positively affected memory acquisition and spatial memory retrieval (Celikyurt et al., 2012). In humans, 14 weeks of lamotrigine treatment in a paediatric BD sample improved working and verbal memory (Pavuluri et al., 2010). In an adult BD sample, Haldane and colleagues (2008) found that 12 weeks of lamotrigine monotherapy had positive effects on working memory, with increased brain activity when performing the N-back task in regions typically activated with this task. Recent work investigating another glutamatergic agent, d-cycloserine (a partial NMDA agonist), in the context of emotional memory found an increase in the specificity of autobiographical memory and the induction of a positive bias in emotional memory categorisation and recall in healthy volunteers (Chen et al., 2021).

To the extent of our knowledge, this is the first time that the effects of lamotrigine are being investigated on emotional memory. Emotional memory is one of the domains most consistently influenced by antidepressant drug action, with this characteristic positive shift being an important mechanism of treatment efficacy (Harmer et al., 2011). The emotional recall task used here allows the exploration of memory for positive and negative self-referential items in the form of a surprise free recall test (Harmer et al., 2011). This task is part of the ETB, which has been extensively used to detect early effects of antidepressant drug action on domains such as perception, categorisation, attention and memory that ultimately affect cognitive and emotional functioning. Interestingly, it has been reported that different types of antidepressants might affect these domains differently (Harmer et al., 2011). In this study, lamotrigine’s effects were restricted to the memory domain, with the rest of the ETB measuring facial expression recognition and attention not yielding treatment-derived effects. This may be due to lamotrigine’s specific influence on the glutamatergic system, known to be involved in memory (Murrough, 2017). With the glutamatergic system receiving increased attention in the pathophysiology and treatment for depressive symptomatology, it is of interest to observe how treatments affecting these systems are effective in doing so. Further research is nonetheless needed to further clarify the effects of lamotrigine on different types of memory.

### Strenghts, limitations and future directions

The present study is the first experimental medicine study investigating the effects of a single oral dose of lamotrigine in healthy volunteers. The use of healthy volunteers allows the characterisation of the effects of a drug on cognitive-emotional functions that are relevant for a disorder, avoiding potential confounding factors such as differences in disorder progression or past treatments. Prior research has highlighted lamotrigine’s positive influence on memory, however the current study explores, for the first time, its influence on the emotional memory domain, which is often impaired in BD.

Although this study has enlightened lamotrigine’s mechanism of action as a mood stabiliser in BD, more specifically in BD depression, it is important to consider these results together with the study limitations. This study primarily found a significant effect on emotional memory (ERC), possibly linked to lamotrigine’s glutamatergic influence. Power limitations may have hindered the detection of further effects on other tasks, like the FERT. Additionally, participants in the lamotrigine group exhibited higher trait anxiety levels, possibly causing an imbalance between groups. This might have affected their self-assessment of side effects, as the groups differed on this measure even at baseline. Nonetheless, despite this potentially negative-valence experience, the lamotrigine group still displayed a positive memory bias.

## Conclusions

This study evaluated the effects of a single oral dose of lamotrigine in healthy volunteers on a range of cognitive-emotional functions using a well validated task battery. Lamotrigine significantly increased the accuracy for the recall of positive vs. negative self-referential descriptors compared to placebo. Although no other effects were found on other subtasks of this battery, this effect is of importance as emotional memory is one of the domains most consistently influenced by antidepressant action in previous studies, and may be closely related to lamotrigine’s glutamatergic effects.

## Data Availability

All data produced in the present study are available upon reasonable request to the authors.

## Acknowledgments

This research study was supported by the NIHR Oxford Health Biomedical Research Centre (OH BRC). The views expressed here are those of the authors and not necessarily those of the NHS, the NIHR or the Department of Health. Liliana Capitão currently works at the Psychology Research Centre (PSI/01662), School of Psychology, University of Minho, supported by the Foundation for Science and Technology (FCT) through the Portuguese State Budget (Ref.: UID/PSI/01662/2020). She is individually funded by a Research Fellowship also from the Foundation for Science and Technology (FCT) (Ref: 2021.00415.CEECIND).

## Ethical standards

The authors assert that all procedures contributing to this work comply with the ethical standards of the relevant national and institutional committees on human experimentation and with the Helsinki Declaration of 1975, as revised in 2008.

## Competing Interests

CJH has received consultancy fees from P1vital Ltd., Janssen Pharmaceuticals, Sage Therapeutics, Pfizer, Zogenix, Compass Pathways, and Lundbeck. CJH holds grant income from Zogenix, UCB Pharma, Pfizer, Janssen Pharmaceuticals. GMG is Chief Medical Officer at Compass pathways, holds shares and share options in Compass pathways PLC and has served as consultant, advisor or CME speaker in the last 3 years for Beckley Psytech, Boehringer Ingelheim, Clerkenwell Health, Compass pathways, Evapharma, Janssen, Lundbeck, Medscape, Novartis, Ocean Neuroscience, P1Vital, Servier, Takeda.

## Author contributions (mandatory) for all authors

TZ was involved in the study conception, data collection, data analysis and manuscript preparation. PAH was involved in data analysis and data checks and prepared the first manuscript draft. LW and AK were involved in data collection and manuscript preparation. GG was involved in the study design, data interpretation and manuscript preparation. LC analysed data, oversaw the study and was involved in data interpretation and manuscript preparation. CJH was involved in the study conception, oversaw the study and was involved in data interpretation and manuscript preparation.

## Notes

### Clinical Trial

NCT04396938

### Author Declarations

Ethical approval was obtained from the Central University Research Ethics Committee (CUREC), from the University of Oxford (UK).

## References

APA. (2022). Bipolar and Related Disorders. In Diagnostic and Statistical Manual of Mental Disorders. American Psychiatric Association Publishing. 10.1176/appi.books.9780890425787.x03_Bipolar_and_Related_Disorders 10.1176/appi.books.9780890425787.x03_Bipolar_and_Related_Disorders

Arnone, D., Horder, J., Cowen, P. J., & Harmer, C. J. (2009). Early effects of mirtazapine on emotional processing. Psychopharmacology (Berl), 203(4), 685–691. 10.1007/s00213-008-1410-6

Bearden, C. E., Glahn, D. C., Monkul, E. S., Barrett, J., Najt, P., Villarreal, V., & Soares, J. C. (2006). Patterns of memory impairment in bipolar disorder and unipolar major depression. Psychiatry Res, 142(2-3), 139–150. 10.1016/j.psychres.2005.08.010

Beck, A. T., Ward, C. H., Mendelson, M., Mock, J., & Erbaugh, J. (1961). An inventory for measuring depression. Arch Gen Psychiatry, 4, 561–571. 10.1001/archpsyc.1961.01710120031004

Bilderbeck, A. C., Reed, Z. E., McMahon, H. C., Atkinson, L. Z., Price, J., Geddes, J. R., Goodwin, G. M., & Harmer, C. J. (2016). Associations between mood instability and emotional processing in a large cohort of bipolar patients. Psychol Med, 46(15), 3151–3160. 10.1017/s003329171600180x

Bogie, B. J. M., Persaud, M. R., Smith, D., Kapczinski, F. P., & Frey, B. N. (2019). Explicit emotional memory biases in mood disorders: A systematic review. Psychiatry Research, 278, 162–172. 10.1016/j.psychres.2019.06.003

Celikyurt, I. K., Ulak, G., Mutlu, O., Akar, F. Y., Erden, F., & Komsuoglu, S. S. (2012). Lamotrigine, a mood stabilizer, may have beneficial effects on memory acquisition and retrieval in mice. Life Sci, 91(25-26), 1270–1274. 10.1016/j.lfs.2012.09.020

Chen, R., Capitão, L. P., Cowen, P. J., & Harmer, C. J. (2021). Effect of the NMDA receptor partial agonist, d-cycloserine, on emotional processing and autobiographical memory. Psychol Med, 51(15), 2657–2665. 10.1017/s0033291720001221

Deckersbach, T., Savage, C. R., Reilly-Harrington, N., Clark, L., Sachs, G., & Rauch, S. L. (2004). Episodic memory impairment in bipolar disorder and obsessive-compulsive disorder: the role of memory strategies. Bipolar Disord, 6(3), 233–244. 10.1111/j.1399-5618.2004.00118.x

Eysenck, H. J., & Eysenck, S. B. G. . (1975). Manual of the Eysenck Personality Questionnaire (junior and adult). Hodder and Stoughton.

Fijtman, A., Bücker, J., Strange, B. A., Martins, D. S., Passos, I. C., Hasse-Sousa, M., Lima, F. M., Kapczinski, F., Yatham, L., & Kauer-Sant’Anna, M. (2020). Emotional memory in bipolar disorder: Impact of multiple episodes and childhood trauma. Journal of Affective Disorders, 260, 206–213. 10.1016/j.jad.2019.09.003

Global Burden of Disease Study, C. (2015). Global, regional, and national incidence, prevalence, and years lived with disability for 301 acute and chronic diseases and injuries in 188 countries, 1990-2013: a systematic analysis for the Global Burden of Disease Study 2013. Lancet, 386(9995), 743–800. 10.1016/S0140-6736(15)60692-4

Godlewska, B. R., & Harmer, C. J. (2021). Cognitive neuropsychological theory of antidepressant action: a modern-day approach to depression and its treatment. Psychopharmacology (Berl), 238(5), 1265–1278. 10.1007/s00213-019-05448-0

Goldsmith, D. R., Wagstaff, A. J., Ibbotson, T., & Perry, C. M. (2003). Lamotrigine: a review of its use in bipolar disorder. Drugs, 63(19), 2029–2050. 10.2165/00003495-200363190-00009

Haldane, M., Jogia, J., Cobb, A., Kozuch, E., Kumari, V., & Frangou, S. (2008). Changes in brain activation during working memory and facial recognition tasks in patients with bipolar disorder with Lamotrigine monotherapy. Eur Neuropsychopharmacol, 18(1), 48–54. 10.1016/j.euroneuro.2007.05.009

Harmer, C. J. (2010). Antidepressant drug action: a neuropsychological perspective. Depression and anxiety, 27(3).

Harmer, C. J., Cowen, P. J., & Goodwin, G. M. (2011). Efficacy markers in depression. J Psychopharmacol, 25(9), 1148–1158. 10.1177/0269881110367722

Harmer, C. J., Duman, R. S., & Cowen, P. J. (2017). How do antidepressants work? New perspectives for refining future treatment approaches. Lancet Psychiatry, 4(5), 409–418. 10.1016/s2215-0366(17)30015-9

Harmer, C. J., O’Sullivan, U., Favaron, E., Massey-Chase, R., Ayres, R., Reinecke, A., Goodwin, G. M., & Cowen, P. J. (2009). Effect of acute antidepressant administration on negative affective bias in depressed patients. Am J Psychiatry, 166(10), 1178–1184. 10.1176/appi.ajp.2009.09020149

Harmer, C. J., Shelley, N. C., Cowen, P. J., & Goodwin, G. M. (2004). Increased positive versus negative affective perception and memory in healthy volunteers following selective serotonin and norepinephrine reuptake inhibition. Am J Psychiatry, 161(7), 1256–1263. 10.1176/appi.ajp.161.7.1256

Hitchcock, C., Newby, J., Timm, E., Howard, R. M., Golden, A. M., Kuyken, W., & Dalgleish, T. (2020). Memory category fluency, memory specificity, and the fading affect bias for positive and negative autobiographical events: Performance on a good day-bad day task in healthy and depressed individuals. J Exp Psychol Gen, 149(1), 198–206. 10.1037/xge0000617

Judd, L. L., Akiskal, H. S., Schettler, P. J., Endicott, J., Maser, J., Solomon, D. A., Leon, A. C., Rice, J. A., & Keller, M. B. (2002). The long-term natural history of the weekly symptomatic status of bipolar I disorder. Arch Gen Psychiatry, 59(6), 530–537. 10.1001/archpsyc.59.6.530

Ketter, T. A., Manji, H. K., & Post, R. M. (2003). Potential mechanisms of action of lamotrigine in the treatment of bipolar disorders. J Clin Psychopharmacol, 23(5), 484–495. 10.1097/01.jcp.0000088915.02635.e8

Lee, C.-Y., Fu, W.-M., Chen, C.-C., Su, M.-J., & Liou, H.-H. (2008). Lamotrigine inhibits postsynaptic AMPA receptor and glutamate release in the dentate gyrus. Epilepsia, 49(5), 888–897. 10.1111/j.1528-1167.2007.01526.x

Malhi, G. S., Mitchell, P. B., & Salim, S. (2003). Bipolar depression: management options. CNS Drugs, 17(1), 9–25. 10.2165/00023210-200317010-00002

Murrough, J. W., Abdallah, C. G., & Mathew, S. J. (2017). Targeting glutamate signalling in depression: progress and prospects. Nat Rev Drug Discov, 16(7), 472–486. 10.1038/nrd.2017.16

Nelson, H., & Willison, J. (1991). The revised national adult reading test–test manual. Windsor, UK: NFER-Nelson, 991, 1–6.

Pavuluri, M. N., Passarotti, A. M., Mohammed, T., Carbray, J. A., & Sweeney, J. A. (2010). Enhanced working and verbal memory after lamotrigine treatment in pediatric bipolar disorder. Bipolar Disord, 12(2), 213–220. 10.1111/j.1399-5618.2010.00792.x

Paykel, E. S. (1987). Cognitive therapy and the emotional disorders: A. T. Beck. The British Journal of Psychiatry, 150(6), 870–871. 10.1192/S0007125000214918

Prica, C., Hascoet, M., & Bourin, M. (2008). Antidepressant-like effect of lamotrigine is reversed by veratrine: a possible role of sodium channels in bipolar depression. Behav Brain Res, 191(1), 49–54. 10.1016/j.bbr.2008.03.007

Prica, C., Hascoet, M., & Bourin, M. (2008). Antidepressant-like effect of lamotrigine is reversed by veratrine: A possible role of sodium channels in bipolar depression. Behavioural Brain Research, 191(1), 49–54. 10.1016/j.bbr.2008.03.007

Sanacora, G., Zarate, C. A., Krystal, J. H., & Manji, H. K. (2008). Targeting the glutamatergic system to develop novel, improved therapeutics for mood disorders. Nat Rev Drug Discov, 7(5), 426–437. 10.1038/nrd2462

Schmidt, K., Cowen, P. J., Harmer, C. J., Tzortzis, G., Errington, S., & Burnet, P. W. (2015). Prebiotic intake reduces the waking cortisol response and alters emotional bias in healthy volunteers. Psychopharmacology (Berl), 232(10), 1793–1801. 10.1007/s00213-014-3810-0

Sills, G. J., & Rogawski, M. A. (2020). Mechanisms of action of currently used antiseizure drugs. Neuropharmacology, 168, 107966. 10.1016/j.neuropharm.2020.107966

Spielberger, C. D., Gorsuch, R. L., Lushene, R., Vagg, P. R., & Jacobs, G. A. (1983). Manual for the State-Trait Anxiety Inventory. Consulting Psychologists Press.

Vajda, F. J., Dodd, S., & Horgan, D. (2013). Lamotrigine in epilepsy, pregnancy and psychiatry--a drug for all seasons? J Clin Neurosci, 20(1), 13–16. 10.1016/j.jocn.2012.05.024

Van der Gucht, E., Morriss, R., Lancaster, G., Kinderman, P., & Bentall, R. P. (2009). Psychological processes in bipolar affective disorder: negative cognitive style and reward processing. Br J Psychiatry, 194(2), 146–151. 10.1192/bjp.bp.107.047894

Warren, M. B., Cowen, P. J., & Harmer, C. J. (2019). Subchronic treatment with St John’s wort produces a positive shift in emotional processing in healthy volunteers. J Psychopharmacol, 33(2), 194–201. 10.1177/0269881118812101

Watson, D., Clark, L. A., & Tellegen, A. (1988). Development and validation of brief measures of positive and negative affect: the PANAS scales. Journal of personality and social psychology, 54(6), 1063.

Young, A. W., Rowland, D., Calder, A. J., Etcoff, N. L., Seth, A., & Perrett, D. I. (1997). Facial expression megamix: tests of dimensional and category accounts of emotion recognition. Cognition, 63(3), 271–313. 10.1016/s0010-0277(97)00003-6

Young, K. D., Bodurka, J., & Drevets, W. C. (2016). Differential neural correlates of autobiographical memory recall in bipolar and unipolar depression. Bipolar Disorders, 18(7), 571–582. 10.1111/bdi.12441

